# Anticipating the novel coronavirus disease (COVID-19) pandemic

**DOI:** 10.1101/2020.04.08.20057430

**Authors:** Taranjot Kaur, Sukanta Sarkar, Sourangsu Chowdhury, Sudipta Kumar Sinha, Mohit Kumar Jolly, Partha Sharathi Dutta

## Abstract

COVID-19 outbreak has been declared as a public health emergency of international concern, and later as a pandemic. In most countries, the COVID-19 incidence curve rises sharply in a short period, suggesting a transition from a disease-free (or low-burden disease) equilibrium state to a sustained infected (or high-burden disease) state. Such a transition is often known to exhibit characteristics of ‘critical slowing down’. Critical slowing down can be, in general, successfully detected using many statistical measures such as variance, lag-1 autocorrelation, density ratio, and skewness. Here, we report an empirical test of this phenomena on the COVID-19 data sets for nine countries, including India, China, and the United States. For most of the data sets, increase in variance and autocorrelation predict the onset of a critical transition. Our analysis suggests two key features in predicting the COVID-19 incidence curve for a specific country: a) the timing of strict social distancing and/or lockdown interventions implemented, and b) the fraction of a nation’s population being affected by COVID-19 at that time. Further, using satellite data of nitrogen dioxide, as an indicator of lockdown efficacy, we find that in countries where the lockdown was implemented early and firmly have been successful in reducing the COVID-19 spread. These results are essential for designing effective strategies to control the spread/resurgence of infectious pandemics.

## I. INTRODUCTION

The outbreak of the COVID-19 disease caused by a novel pathogenic coronavirus (SARS-CoV-2) - which initiated in Wuhan, China in December 2019 - is a global challenge for the healthcare, economy and the society [1]. The World Health Organization (WHO) assessed the massive epidemics of the disease (COVID-19) and declared it as a Public Health Emergency of International Concern (PHEIC) [2]. Since the Wuhan outbreak, nearly all the United Nations member countries have experienced a rapid spread of the virus and are taking preventive measures to overcome the threats posed by the pandemic [3]. Over the past years, several waves of viruses such as influenza, cholera, and HIV have transmitted across the world and caused huge threat to human health. Investigations on the interventions of these outbreaks have given rise in predictive theory of infectious diseases. Importantly, prior understanding of the epidemic spreading of COVID-19 can provide effective mitigation policy.

The COVID-19 disease can spread in a population through infected symptomatic/asymptomatic individuals who come in contact directly/indirectly [4]. Thus, concerned with the public health and well-being affected due to COVID-19, various countries have adopted comprehensive clinical and non-pharmaceutical strategies. The non-pharmaceutical interventions are converging to social distancing, such as the closure of schools, ban of large gatherings, isolation of symptomatic individuals, and monitoring travelers, particularly to those from COVID-19 hotspots [5–8]. There also exists evidence of similar non-pharmaceutical interventions to mitigate the 1918 influenza pandemic [9, 10]. Evidences also highlight the importance of mitigation interventions in controlling the transmission of the SARS-CoV-2 virus [6, 11, 12]. Nonetheless, the timing of the implementation of strategies vary between countries and can significantly influence the incidence curve of the epidemic [13].

The COVID-19 incidence curve of total confirmed cases for many countries initially demonstrates a gradual increase near the start of the epidemic and is often followed by a sudden shoot or a transition to a supercritical state [14–18], as the disease spreads (major outbreak due to human-to-human transmission). This sudden transition places a considerable burden on the limited availability of the public health resources required to treat the disease and inhibit its further spread. Most of the studies on sudden transitions concern with catastrophic shifts associated with a saddle-node bifurcation, however epidemic transitions are non-catastrophic and associated with a transcritical bifurcation [17, 19]. In general, an epidemic transition occurs when the basic reproduction number (or 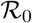) of the disease becomes greater than one and a population moves from a subcritical to a super-critical state. However, in COVID-19, in many countries major outbreaks do not occur initially, though the 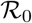 of the disease is known to be more than one from the very beginning [20]. In fact, this may be associated with a tipping delay where a population faces the first major outbreak at a higher value of 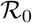 than one [14, 21], impending our ability to mitigate. Thus, it is crucial to anticipate this precarious transition to take effective controlling measures for the outbreak. There exists a rich history of investigations that can predict processes which could lead to ecological outbreaks [19, 22–24]. Theory suggests applicability of a variety of leading generic indicators, widely known as Early Warning Signals (EWSs) (e.g. variance, autocorrelation, skewness, and kurtosis) to identify the proximity of a system to such a critical transition [22, 23, 25, 26]. For instance, in time-series data following ancient abrupt climate shifts, EWSs can be identified before the critical transition took place [27]. Similarly, EWSs were seen in the resurgence of malaria in Kericho, Kenya [18].

EWSs are hallmarks of critical slowing down (CSD) of a system as it approaches a catastrophic/non-catastrophic transition. The phenomenon of CSD owes to the loss of resilience in the system such that even small disturbances can invoke an often irreversible transition to an alternative stable state [28–31]. In particular, dynamical systems are continuously subject to shocks which may be extrinsic or intrinsic perturbations. In epidemiological theory, intrinsic perturbations can be determined by the pathogens novelty in a new host which may depend upon various health factors associated with the host. Further, the mode of transmission, person-to-person contact, and number of imported cases may account for external perturbations for disease spread. Increased perturbations may drive a system far from its original state and can increase the time required for fluctuations in the number of cases to dampen. Thus, the system loses its resilience as it may eventually diverge at a transition from a low burdened to a high burdened state. The phenomenon of CSD can be captured as a large time taken by a system to return to its previous states due to which the rate of return of a system decreases prior to a transition. Moreover, it leads to an increase in the short term memory of a system, this feature can be identified by the changes in the correlation structure of a time-series preceding a critical transition [22, 23, 26, 32, 33].

Model based epidemiological investigations predict the phenomenon of CSD preceded by the epidemic transitions [15, 34, 35]. These studies are build on the applicability of CSD-based EWSs to anticipate disease emergence. However, construction of emerging disease models can be complicated partly due to non-linearity in many natural systems. Also data availability of key epidemiological parameters such as rate and mode of transmission, duration of infection, novelty of the pathogen in a new host can pose a barrier towards disease predictive theory. The key support of CSD-based EWSs analyses over modelling prediction is that it does not require comprehensive data calibration and can be calculated using observed data. Further, it is studied that imperfections in the disease data does not form a barrier in applicability of EWSs [15].

To mitigate the epidemic, China strictly restricted public movement and followed measures of quarantine and symptomatic isolation 24 days later (i.e. 23 January) to the arrival of the first reported case. The total reported cases (confirmed) at the time of the lockdown were nearly 623 (accounting for approximately 4.4732 × 10^−7^ of the total population). The daily increase in the number of confirmed cases in China, thus, saturated nearly in mid of March, hence flattening the incidence curve of the total confirmed cases. European countries adopted different non-pharmaceutical measures to intervene in the disease transmission. The spread began later in Italy as compared to China; however, the strict interventions were initiated on 9 March, which marks a gap of nearly 40 days from the first reported case with ≈ 1.22 × 10^−4^ proportion of the cases. Spain, which is continued to suffer severely by the virus, reported its first infected case on 1 February and took nearly 45 days when the proportion of affected cases was more than 9.05 × 10^−5^, to put the country on lockdown (see *SI Appendix, Table S1*). India confirmed its first case on 30 January and prompted “Janata curfew” on 22 March, followed by a nationwide lockdown on 25 March for complete cessation of public contacts (nearly 55 days after the first case being reported), however the proportion of cases were approximately 2.36 × 10^−7^ of its population as COVID19 infected cases, while this proportion was more than 1.7 × 10^−4^ in the US. Therefore, it is essential to unfold how prolonged gaps between the arrival of the epidemic and non-pharmaceutical interventions such as quarantining/social distancing can influence public health and the environment at a national as well as a global scale. More interesting is to understand if the EWSs can be useful to stifle the spread of an epidemic.

In this work, we analyse how the timing of strict controlling strategies influence the COVID-19 incidence curve of the total confirmed cases in different countries. We first use the ‘change in the gradient’ analysis (for details see *SI Appendix, section 4.: Detection of the transition phase*), to estimate the emergence of the transition phase in incidence curves. The occurrence of CSD is then analysed using the data prior to the transition. We calculate the variance and lag-1 autocorrelation function of the time-series data of the cumulative confirmed cases each in nine different countries. Our work suggests that the dynamics of incidence curve in the initial days (depending upon the country), since the first reported case, can signal an upcoming sudden rise in the cumulative number of infected cases. Thus, preliminary interventions are crucial for an effective and timely containment of the disease emergence or resurgence. Delay in the strict surveillance and control measures can increase the time to contain the spread, which in turn will affect a larger proportion of the population. Furthermore, the proportion of the affected cases on the commencement of public health measures plays a significant role in containing the epidemic in each country. The time gap of implementation of interventions from the arrival of the first case is almost similar for many countries such as Italy, India and Germany. However, the EWSs depict an upcoming rise in Italy and Germany relatively earlier than in India. The relatively low proportion of the affected cases in the case of India compared to Italy or Germany can be a significant factor explaining this slow rise for India but a relatively disruptive situation in the other countries. Thus, a combination of these two factors for India may restrict the extent of COVID-19 spread in the country, as compared to many other countries across the world. Importantly, despite of controlled situation upto 29 April, India, having greater carrying capacity for the disease and challenging sanitization control [37], needs strict and highly effective interventions for continued suppression in the daily number of cases. We conclude that model-independent forecasting systems can be applied to clinical data sets for predictability of the disease re-occurrence and formulate control policies.

## II. RESULTS

We obtain the data sets of the cumulative number of the COVID-19 cases from the date of reporting of the first affected person up to 29 April, 2020 each for India, China, South Korea, the United States (US), Singapore, Germany, Italy, United Kingdom (UK) and Spain (for the data source see *Materials and Methods*). Figure 1 depicts the incidence curve of the affected population in each of these countries. Interestingly, it is noted that the incidence curve of the confirmed cases follows a slow increment during initial time period ranging from ≈ 20-50 days, for different countries, which can be interpreted as a time window to control the epidemic promptly and effectively. Since human to human contact is a leading transmitter of the disease, therefore, by-passing a certain threshold of infected cases, the incidence curve shows an increasing slope and finally depicts a transition in the number of infected cases (see Fig. 1) [38]. It is important to note that the growth in number of cases for China and South Korea, the countries which initiated public monitoring/social distancing actions relatively earlier as compared to the other countries, saturates after nearly 3-4 weeks from the initiation of the lockdown. The shift of the COVID-19 from a low-burden to a high-burden state can be associated with the phenomenon of critical transition. Thus we employ statistical methods that can monitor the onset of the transition phase and provide insights into the incidence curve so as to suggest establishing worldwide disease elimination campaigns.

**FIG. 1.**
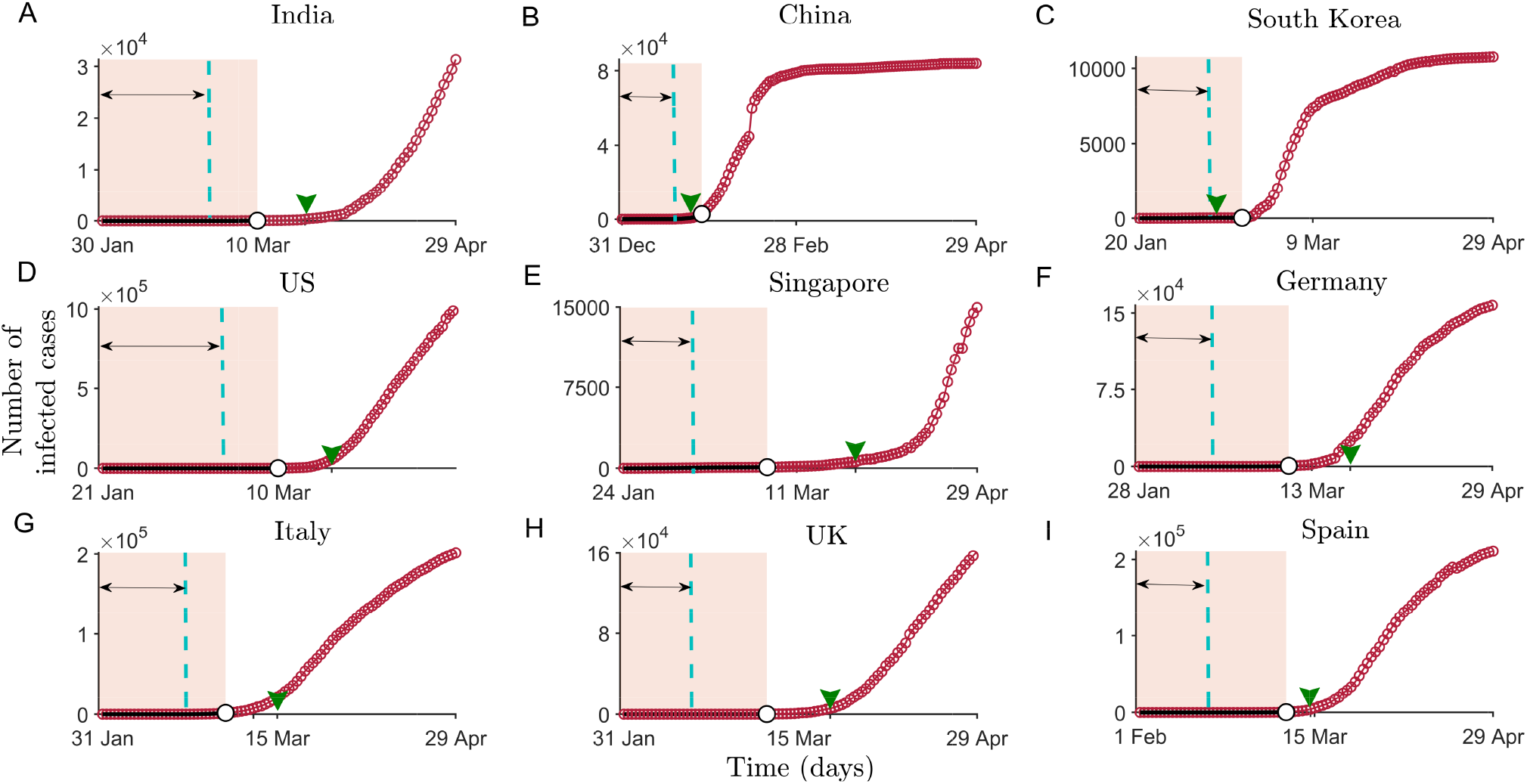
Time-series constructed as cumulative number of infected cases in nine different countries across the world, from the onset of the epidemic in the respective countries up to 29 April. Shaded regions depict the pre-transition phase for different countries from the onset and marks the data used to compute the indicators of critical slowing down. The double sided arrow marks the size of the moving window (up to the vertical dashed line). In the subfigures, the arrow-head on the *x*-axis marks the beginning of the officially recorded social-distancing and/or lockdown dates (see *SI Appendix, Table S1*).

### Signals of critical slowing down

To estimate statistical indicators anticipating the upcoming shifts in each country, we consider the data of cumulative daily number of COVID-19 cases before a transition is detected in the incidence curve of the epidemic (shaded regions in Fig. 1) (for most of the countries a transition threshold is detected by a gradient change analysis, for details see *SI Appendix, section 4.: Detection of the Transition Threshold*). To examine whether the system slows down to recover from perturbation while approaching the transition, we calculate the variance and autocorrelation at first lag (ACF(1)) of each extracted data for all the nine countries (see *Materials and Methods*). We have also calculated few other generic EWSs of CSD, like density ratio, skewness and kurtosis (for details see *SI Appendix, section 1.: Early Warning Indicators*). CSD is reflected in systems near a critical transition through an increase in the variance and autocorrelation. We observe that the short term memory of the time-series data exhibits an increasing trend in most of the countries (Fig. 2). However, there are no positive signals of CSD exhibited by ACF(1) for the data sets of India as well as Italy (Figs. 2J and 2P). The increase in the variance forewarns a sudden rise in the number of the COVID-19 cases for these countries. Furthermore, the strength of the signals vary amongst countries depending upon the data sets determining the cumulative number of affected population in individual countries. For instance, we observe a weak increase in variance in case of Singapore, and the trends in China and the US are observed to be very strong, with ACF(1) approaching close to 1 (see Fig. 2K and 2M) [32]. Since the time lag of up to almost two weeks is expected for the detection of symptomatic cases [39], the analyses suggests that the total cases gathered when the phenomenon of CSD is observed must be infected with the disease around two weeks ago. Thus, an early preventive and surveillance strategies can be capable of suppressing the severity of COVID-19 outbreak [40].

**FIG. 2.**
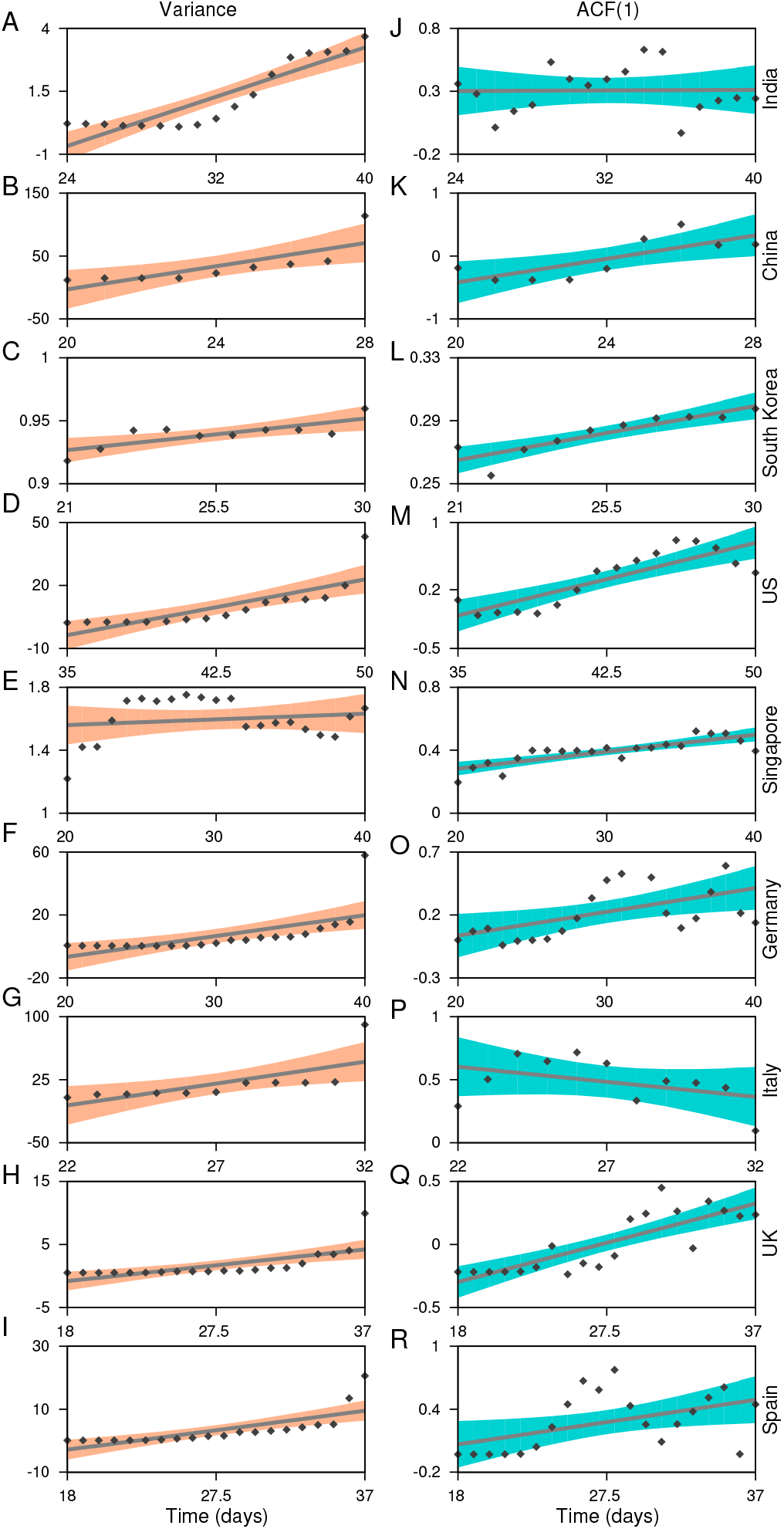
Statistical estimates used to analyse the signals of a forthcoming transition in the COVID-19 incidence curve. Figures on the left panel depicts the variance each for India, China, South Korea, USA, Singapore, Germany, Italy, UK, and Spain. The right panel shows the lag-1 autocorrelation of the time-series data analysed in the corresponding countries. Scattered points are the estimated values of the respective slowing down indicators. Solid lines reflect the increasing/decreasing trend in the indicators and are obtained by fitting linear regression models. The shaded regions are the confidence bounds for the fitted models.

### EWSs and enforcement of interventions

The timing of intervention measures varies among the countries. Notice that, except China and South Korea, in other countries EWS analyses are carried out using the data before the implementation of social distancing measures. China was the first country to take the containment measures, nearly 24 days after the beginning of the epidemic, while Italy took around 40 days, and other countries followed afterwards. As a consequence, the COVID-19 incidence curve in China flattened nearly after 20-25 days of implementing the intervention measures. Similar to China, South Korea adopted different combinations of controlling measures around mid February (in the time window of 20-25 days since the epidemic began there). This measure was accompanied by a drop in the number of cases, and the curve followed the pattern as observed for China (Fig. 1 C). The rising indicators of CSD also suggest that the time gap in implementing the protocols such as the closure of public gatherings, controlled public movement, lockdown can significantly influence the incidence curve and result in the extended time required to flatten it. However, the interventions around 2-3 weeks prior to the change in the correlation pattern as well as variance in each of these countries can slowly hamper the daily increase in the number of cases.

The scenario is quite different in the case of India. The EWSs weakly signal the behavior of CSD within the initial 40 days of the disease emergence (Figs. 2A and 2J). Due to a rise in the number of daily cases, we also analyse the EWSs in the incidence curve for India considering the cumulative number of infected cases of upto the beginning of the nationwide lockdown (25 March, Fig. 3A) since the reporting of the first case. Here, we observe increasing trend in each of the generic indicators capable to capture the phenomenon of CSD. The variance, autocorrelation, skewness and kurtosis captures the strong signals of CSD (Fig. 3B-3F).

**FIG. 3.**
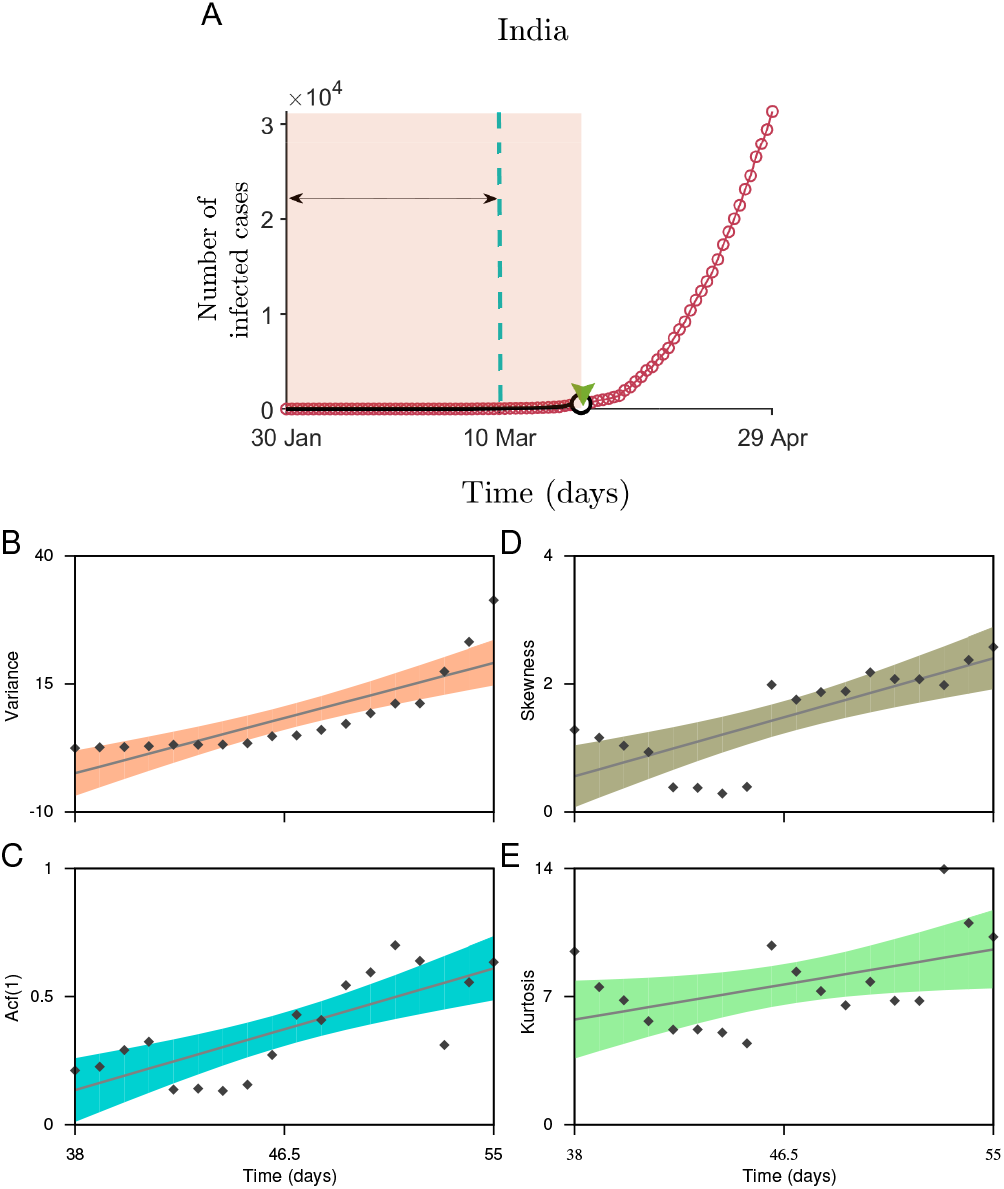
Statistical analysis to measure the indicators of CSD for the dataset the of India upto the date of the nationwide lockdown. A: The incidence curve depicting the fraction of people infected from 30 January up to 29 April. The shaded region is the data used to calculate the changes in the generic indicators. The arrow marks the size of the rolling window used to calculate the statistical signals. The estimated values of the slowing down indicators B: variance, C: autocorrelation function at lag-1, D: skewness and E: kurtosis. Solid lines are the fitted linear regression models to analyse the trend in the indicators along with the confidence bounds (shaded regions).

### Onset of social distancing practices and the affected population density

Another important aspect is to consider the reported proportion of a population affected at the time of the implementation of intervention measures. So far, Germany which accounted for one of the largest outbreaks in Europe around mid of the March, had visible signals of the forthcoming transition (Figs. 2F and 2O). It is noted that each of the countries, namely India, Germany, and Italy, adopted concerned public health measures around the time when the EWSs were visible in their respective data sets (see the arrow-heads on the *x*-axis in Fig. 1). However, the fraction of the population affected by the time in Germany and Italy was much higher (approximately 2.9 × 10^−4^ and 1.2 × 10^−4^, respectively) as compared to that for India − 2.36×10^−7^. Thus, the incidence curve projected a significant rise in these two countries, whereas the rise in the number of cases in India is relatively slow and is expected to follow a similar response owing to the effectiveness of these interventions. Overall, our analyses suggest that delayed interventions (depending upon the signals of CSD) along with the fraction of the affected population can influence the country-wise variation in the daily number of rising cases.

### Sensitivity analysis of the generic indicators

The choices made to remove/filter out nonstationarities in the time series datasets using Gaussian detrending can also influence the trends observed. Thus, it is necessary to test the robustness of the estimated trends towards the choice of rolling window size and the filtering bandwidth. Here, we employ sensitivity analysis for the variance (see Fig. 4) and ACF(1) (see Fig. 5) using the CSD data set. Sensitivity analyses ease outs to disentangle accurate signals of an impending transition from the false ones for a wide range of window sizes and bandwidths. We use Kendall-*τ* estimates of these indicators for all the combinations of these two parameters (for details, see *Materials and Methods*). Furthermore, we test the sensitivity of these parameters on the P values of the estimated indicators (for details see *SI Appendix, section 3.: Sensitivity analysis*).

**FIG. 4.**
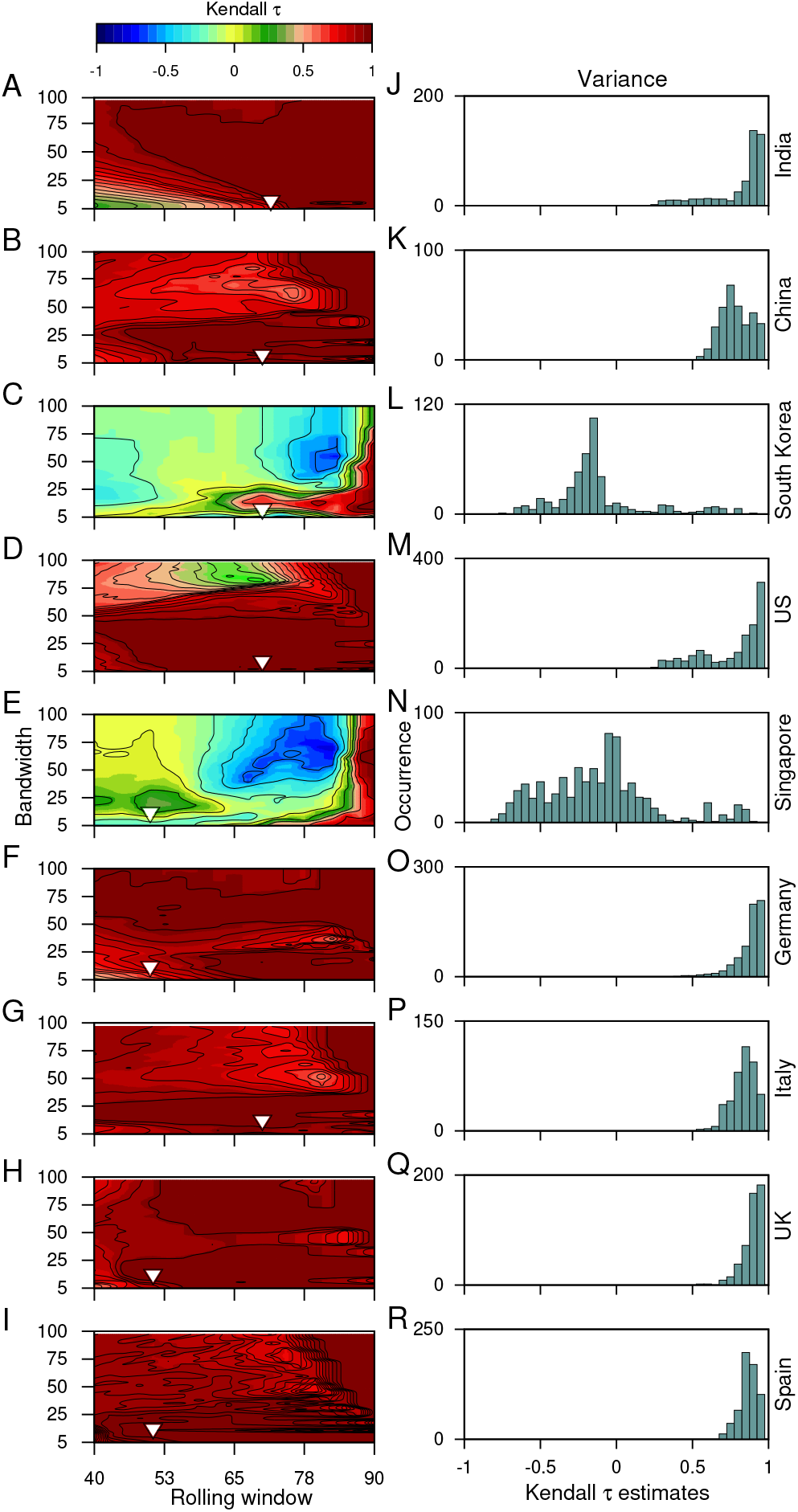
The sensitivity of the choice of the rolling window size and the filtering bandwidth to estimate the EWSs. Contour plots demonstrate the effect of moving window size and the filtering bandwidth on the trends observed while calculating the changes in the variance of the time series, using the Kendall-*τ* test statistic. Panels on right show the frequency distribution of the trend statistic. The inverted arrows mark the choice of the filtering bandwidth and moving window size used to capture the trends in the variance of the time series data.

**FIG. 5.**
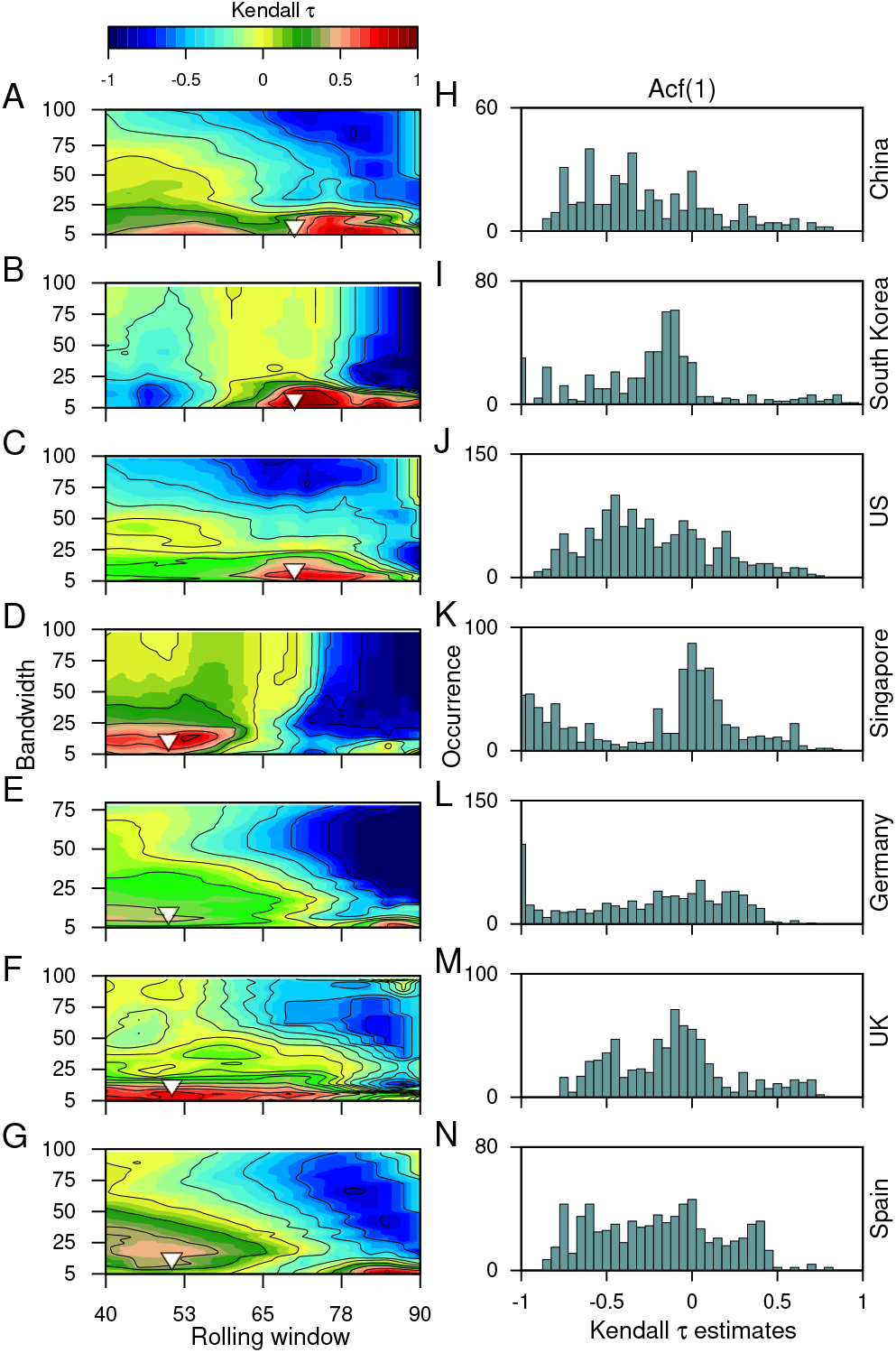
The sensitivity of the choice of the rolling window size and the filtering bandwidth to estimate the EWSs. Contour plots demonstrate the effect of moving window size and the filtering bandwidth on the trends observed while calculating the short term correlation pattern using ACF(1) of the time series data, using the Kendall-*τ* test statistic. Panels on right show the frequency distribution of the trend statistic. The inverted triangles mark the choice of the filtering bandwidth and moving window size used to capture the trends in the ACF(1) of the time series data.

We find that the observed trends in the variance are robust to the choice of parameters and does not vary between the data sets of most countries. High bandwidths reveal the opposite outcome of the variance for the data sets of South Korea (Figs. 4C and 4L) and Singapore (Figs. 4E and 4F). Since we use the bandwidth, which gives the best fit and does not over-fit or under-fit the data therefore, the choice of window size can influence the observed trends. In our work, we find a large size of the rolling window can alter the EWSs analysis and produce misleading estimates for the autocorrelation function (Figs. 5A–5N). False signals of an alarming situation can deviate from understanding the gravity of any situation and intensity of surveillance needed. Thus, the choice of these parameters is crucial in anticipating the signals of a forthcoming transition and implementing convincing public health measures.

### Surrogate analysis

The lower number of data points available for the analysis can lead to feeble trends and influence the probability of occurrence of the increased signals of CSD by chance. Further, due to undocumented patients, there is always a chance of stochasticity in the number of reported cases. Thus, we studied the likelihood of coincidence in the occurrence of trends in the variance and the ACF(1) observed in our original data sets by investigating the indicators in the surrogate time-series (see *Materials and Methods*). The surrogate time-series is generated to follow similar distribution (mean and variance) of the data time-series before the episode of a sudden rise in the number, denoted by shaded regions in Fig. 1 (see *Materials and Methods*). Figure 6 depicts the distribution of the test statistic of the surrogate time-series. Solid lines show the trend estimate obtained for the original time-series. We calculate the probability of randomness of our observed estimates as the fraction of 1000 surrogate time series having trend statistic of same or higher values than the original trend, i.e., *P* (*τ*^∗^ ≤ *τ*). The probability to, by chance, obtain similar trend statistic varies from country to country, depicting significant estimates for changes in the variance, except for South Korea (Figs. 6B and 6I) and Singapore (Figs. 6D and 6K). While in the case of the US, the probability of randomness in our observed estimates is very less (Figs. 6C and 6J), rapid spreading in the epidemic makes it keystone to consider applicability of EWSs to warn-off such events. The probability estimates *P* obtained by bootstrapping the data sets for each of the countries are given in the *Table I*.

**FIG. 6.**
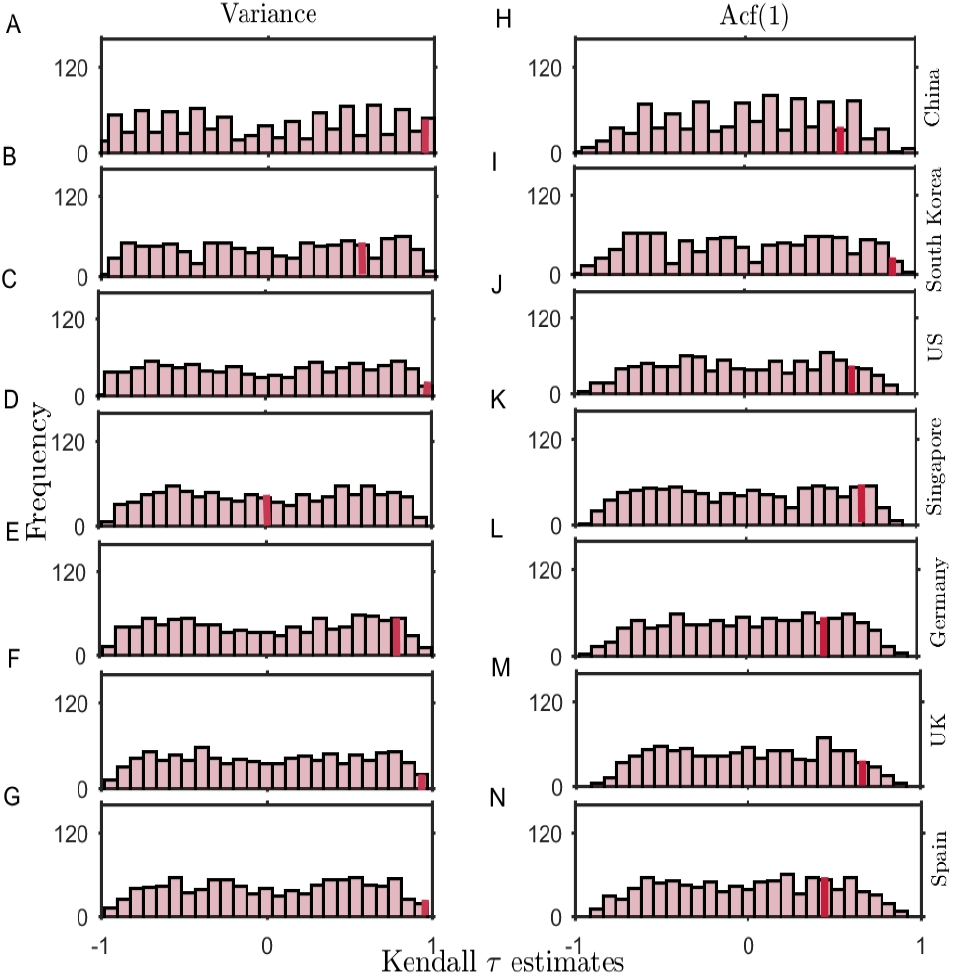
The probability distribution of Kendall-*τ* test statistic on a set of 1000 surrogate time-series generated by bootstrapping and shuffling (with replacement) the residual time-series of the original data. Histograms depict the distribution of the test statistic for the surrogate time-series variance (left panels) and autocorrelation function at lag-1 (right panels). Solid lines indicate the limit beyond which the Kendall-*τ* of the surrogate data is higher than the statistic observed in the ACF(1) of the original time-series.

**TABLE I.** Probability of, by chance, obtaining the observed trend statistic of the original data for the set of 1000 surrogates having similar distribution (mean and variance) as the original data-sets. The likelihood of randomness in the estimated variance and ACF(1) is mentioned for the data-sets of each country studied in the work.

| Country | Kendall-τ<br>(variance) | Kendall-τ<br>(ACF(1)) |
| --- | --- | --- |
| China | 0.04 | 0.09 |
| South Korea | 0.21 | 0.01 |
| US | 0.001 | 0.1 |
| Singapore | 0.48 | 0.09 |
| Germany | 0.08 | 0.20 |
| UK | 0.001 | 0.05 |
| Spain | 0.001 | 0.21 |

Overall, we find a low probability of randomness in both the ACF(1) and the variance estimates for most of the cases. However, the observations are more significant for the variance. This analysis suggests the robustness of the variance as an EWS in predicting the signals of CSD.

### Impact of COVID-19 spread on the atmospheric total-column NO_2_ density

The rigor of social distancing/intervention strategies can be measured by atmospheric data, as the lockdown periods have witnessed better air quality across the globe [41]. We note that anthropogenic NO_2_ is emitted pre-dominantly at the surface from transportation activities, industries and power plants. NO_2_ emitted has a short lifetime and can be transported upto a few hundred metres during the day. Therefore, NO_2_ is expected to be a profound indicator of the efficiency of lockdown measures enforced by the countries. Thus, we first obtain time-series of triads of the population-weighted total-column NO_2_ (molecules/cm^2^) density over the length of the study period for the nine countries considered in this work (the circular points in Fig. 7) (for the NO_2_ data source see *Materials and Methods*). The solid curve in each subfigure of Fig. 7 represents a 10-triad moving window average. In the majority of the countries, the timing of NO_2_ decline concurs with the spread of the virus and the onset of pragmatic lockdown in a country may be hypothesized by the reversal (or break) in the trend of NO_2_. In China (Fig 7B), the decreasing trend in NO_2_ is evident from January end till February; after that, it starts increasing which is coincident with the dynamics of the spread of COVID-19 disease. In India (Fig 7A), South Korea (Fig 7C), US (Fig 7D), Italy (Fig 7G) and Spain (Fig 7F), the decreasing trend in NO_2_ coincides with time of the rapid spread in the virus (Fig. 1). We estimate that after the date of official enforcement of lockdown, the time-averaged NO_2_ decreased by 26.6% in China and 55.6% in Italy compared to the pre-lockdown period. Spain, USA and India have also seen a significant decrease after the lockdown was enforced in these countries by 33%, 22.9% and 11.8%, respectively. Whereas, it increased in UK and Germany by 18% and 32%, respectively, even after the initiation of lockdown which indicates an inefficient closure of anthropogenic activities (like road and rail transport, industries and power plants). The spatial distribution of total-column NO_2_ for all the triads starting 28 Dec, 2019 up to 10 May, 2020 can be visualised in *SI Appendix, Movie 1*. It should be noted that we did not control for meteorological variations which may have a significant impact on total-column NO_2_ over the period of our study [42]. Overall, amidst the fears of the novel coronavirus, the countries where the lockdown intervened are expecting a rejuvenated environment. However, at the same time, possibilities to lower down air pollutants when the world is not facing such harsh conditions is also important to understand.

**FIG. 7.**
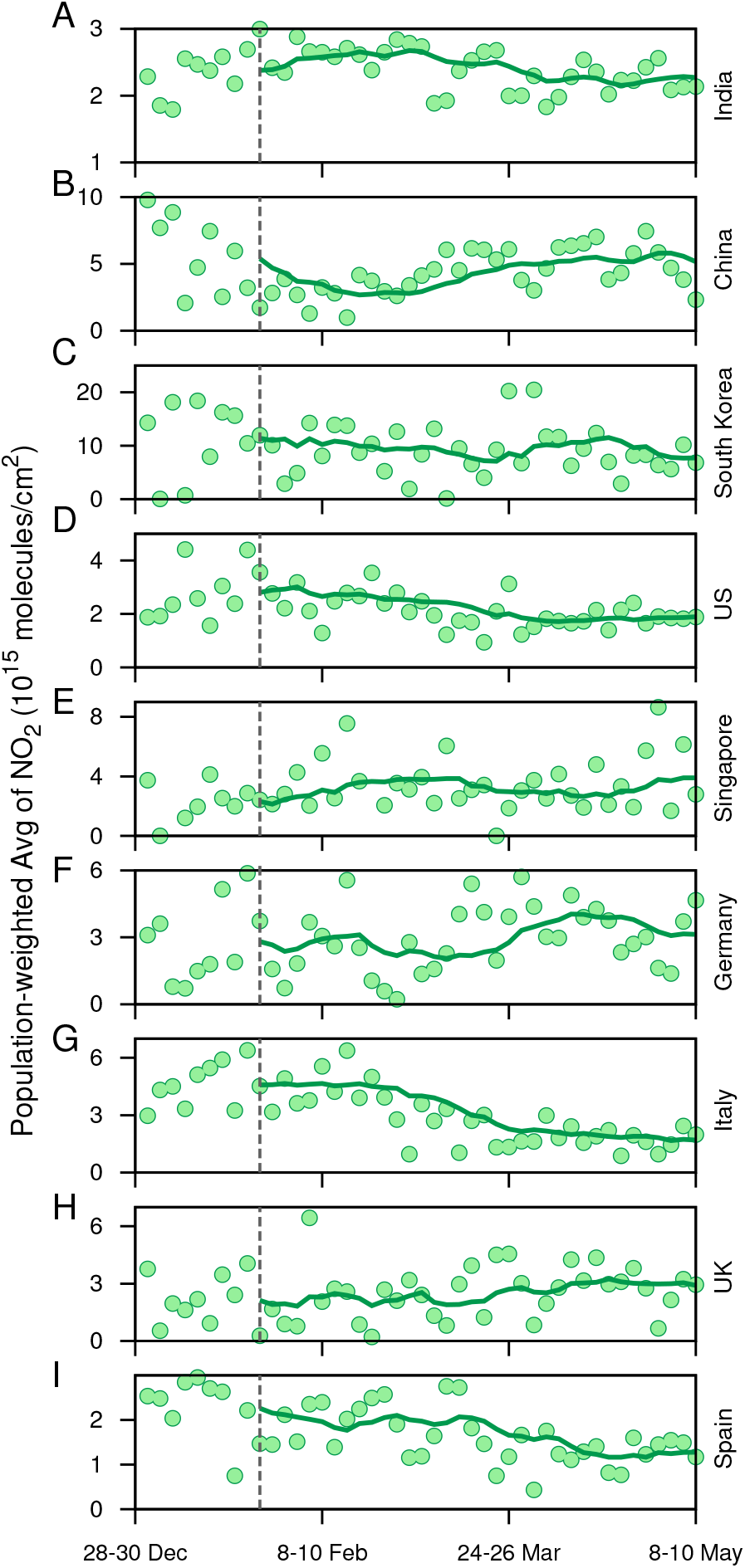
Time-series of triads of the population-weighted total-column NO_2_ (molecules/cm^2^) density over the length of the study period for the nine countries considered in this work (depicted by the circular points). The solid curve in each subfigure represents a 10-triad moving window average of the time-series.

### A minimal stochastic model

We propose a minimal kinetic model for the short term prediction of the spreading of COVID-19 disease. Suppose that the only processes are infection and recovery. The processes can be described as:

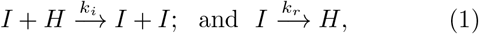

where *I* and *H* are infected and healthy people, respectively, and *k_i_* and *k_r_* are rate constants for infection and recovery. The first equation shows that if *I* is the infected people, then *H* becomes *I* at a rate *k_i_*; and the second equation indicates that *I* recovers at a rate *k_r_*. A minimal kinetic model can be formulated as ordinary differential equations for the population of *I*, as:

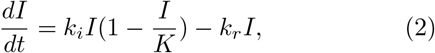

where *K* is the size of the population.

We develop a master equation for the infected population by considering the two elementary processes (Eq. 1). The transition probability at which the number of infected population increases from *i* to (*i*+1) is w(*i*+1|*i*)= *k_i_i*(1−*i*/*K*), and the rate at which the number of infected population reduces from *i* to (*i*-1) is w(*i* − 1|*i*)= *k_r_i*. From these, the probability of finding *i* infectives in the system at time *t*, *P* (*i*, *t*) can be obtained from the following equation:

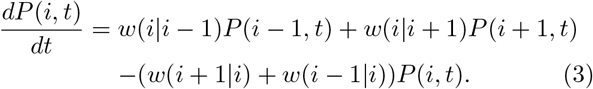

The above probabilistic model is solved by the kinetic Monte Carlo simulations by means of the Gillespie algorithm, which incorporates the intrinsic noise [43]. The algorithm considers each of the events as individual realisations of Markov process. The time and species numbers are updated stochastically by choosing the random processes.

To simulate the system (Eq. 3), we first obtain the parameters from the cumulative time series data of confirmed cases for India, China, and South Korea. In the data sets, we fitted the below logistic function (which is a solution of Eq. 2):

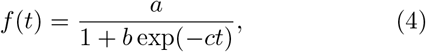

where *a*, *b*, and *c* are parameters. Once we obtain these parameters for an individual country, we map them to our model and find the system parameters *k_i_*, *k_r_* and *K*, and *i*_0_ is the initial infected population. We list those parameters below:

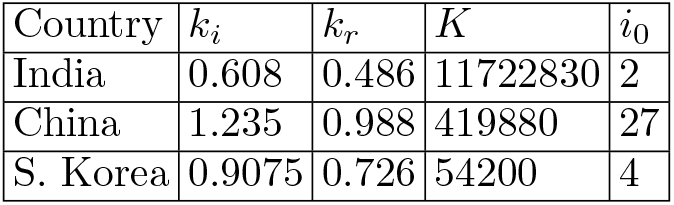

Then the above parameters are used to solve the Master equation (Eq. 3), and we perform Monte-Carlo simulation to get stochastic trajectories up to 15 April. We present the simulated stochastic trajectories in Fig. 8. For each country, we have five trajectories. For China and South Korea, we find that our stochastic trajectories are consistent with the real time-series of the number of infected people. However, for India, our result shows that on 20th May 2020, the number of infected people can go up to approximately 1,09,262 (which is an average of final values of the five simulated trajectories).

**FIG. 8.**
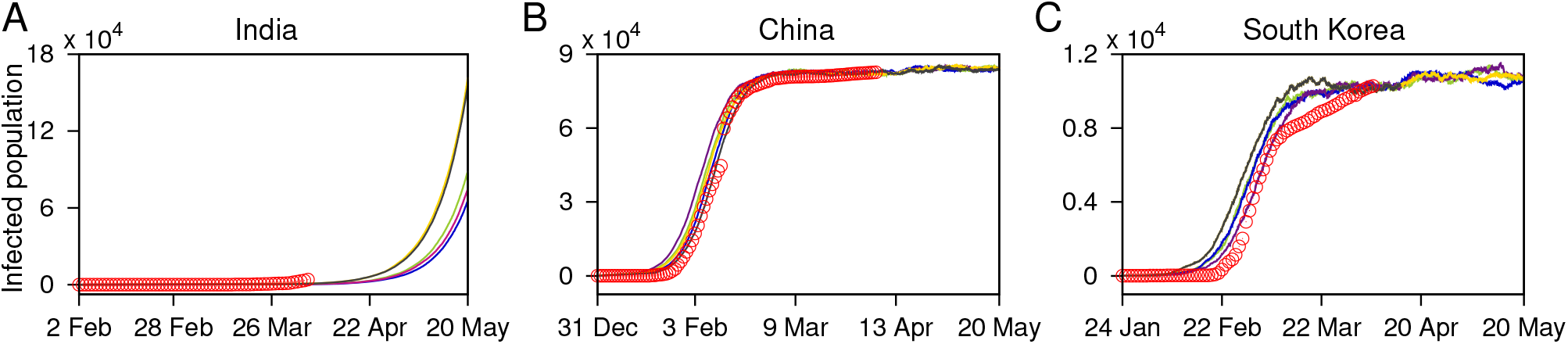
Stochastic trajectories (marked with pink, yellow, grey, green, and blue curves) of the infected population generated using the Gillespie algorithm for: A. India, B. China, and C. South Korea. The original data sets up to 6 April for the respective countries are depicted by circular (red) points, for a comparison.

The problem of predicting the spreading of COVID-19 is a complex one and depends on many factors like social distancing, an early detection of the disease, the detection of major hubs of the disease, etc. Here, we have provided a minimal kinetic model which uses the trends of the available data and may work only for short term prediction.

## III. DISCUSSION

The COVID-19 pandemic revealed an exponential rise in the reported number of cases and affected the public health ranging from mild to severe conditions. Countries across the world are combating the spread of the coronavirus through various social distancing/intervention measures such as closure of schools and universities, banning of public events and large gatherings, isolation of symptomatic COVID-19 cases, mass quarantines etc. For national as well as international control of public health, it is crucial to understand the significance of the onset timing of such measures [44].

The World Health Organisation lately reports new cases being detected in several new countries across the globe [45–47]. Our study can provide an insight to tackle the ongoing pandemic and its associated incidence curve in the context of the timing and strength of the interventions. We use the data of the number of COVID-19 cases in nine different countries to investigate some statistical patterns in the incidence curves. The number of cases covers a small fraction of the population during the initiation of the epidemic, and the fraction remains nearly stagnant ranging nearly from 20 − 50 days from the arrival of the first case. Further, the number of cases start increasing rapidly, and in a relatively shorter span a significant fraction of the population can be affected. This trend is analogous to the idea that the incidence curve remains close to one stable state for sufficient time and, crossing a time threshold invokes a sudden shift/transition to another stable state, where a significant fraction of the population gets affected. In our work, we employ statistical indicators of critical slowing down to check if such transitions can be signalled beforehand and how the anticipation of such transitions can help mitigate such crisis at a policy level.

We observe that the initial time window from the arrival of the first case in each country signalled an impending transition. An increase in the ACF(1) of the data as well as variance, before an actual rise in the number of cases indicates the phenomenon of critical slowing down. Our work suggests that while non-pharmaceutical interventions are necessary to mitigate such epidemic, the timing of initiation of concerned actions can strongly influence the outcome of the situation. Owing to the time lag in the detection of symptomatic cases, the statistical indicators suggest that the time period of 2-3 weeks before an impending transition being crucial to suppress the loss of public health. The controlled response of the epidemic incidence curve for China and South Korea can be associated with the time distance between implementation of interventions and the transition point. Both these countries initiated interventions before the visible signals of CSD in the incidence curve. Thus timely interventions played one of the important factors to suppress the fluctuations in the number of cases and shape the curve. Thus, EWSs analysis is crucial while defining the onset of the interventions and suppress the rise of daily cases. Importantly, another crucial aspect is the proportion of affected cases in each country, i.e., a measure of the fraction of the country’s population, and not the absolute numbers, which is infected at the time of interventions such as a strict lockdown. As probability of the propagation of disease can be thought of as mostly similar or equal amongst individuals across the globe, it depends upon the fraction of infected cases in each country during the beginning of interventions. For instance, the EWS analyses anticipated the upcoming rise in the incidence curves for both India as well as Italy, and interestingly, both the countries imposed individual nationwide lockdown near the situation close to the transition (see *SI Appendix, Table S1*). However, the control in India depicts better results in altering the incidence curve than that in Italy. The alterations in the incidence curve is most likely to be a consequence of a difference in the proportion of cases affected by the epidemic at the beginning of mitigation strategies. India resembling China in terms of the total population density accounted for approximately 2.36 × 10^−7^ cases of the total population, while Italy with relatively less population density crossed 1.22×10^−4^ cases of their total population. Thus, even with imposition of the public health measures near the signals of CSD, the outcome for both the countries can dramatically vary. The variation is the consequence of the proportion of affected cases when visible signals of EWSs are observed and at the time of interventions. This suggests that the proportion of affected population during visible signals of CSD is key to shape the disease incidence curve. The strength of the signals can alter the duration and scale of the interventions needed. Furthermore, the disruptive situation in the US is indicated by EWSs, as the EWSs indicators show significant trends for the US, on addition with the large fraction of population being affected at that time. Thus, sharp rise in the number of cases for the country is a consequence of both the delay in effective social distancing interventions as well as a large proportion of affected cases at that time. Overall, our work suggests that in almost all the countries, a sharp upcoming rise in the incidence curve can be captured using statistical measures, before the actual transition.

Another horizon the infectious coronavirus puts forward is on the quality of air pollution in countries where social distancing/lockdown is enforced. NO_2_, which is majorly emitted from anthropogenic activities like land transportation, industries, and energy sectors, was estimated to decrease in consequence to lockdown measures implemented by the government of respective countries. Population weighted average column NO_2_ was found to decrease with amplification in a number of cases across most of the countries. Apart from this, NO_2_ column quantities may be used as a proxy to estimate the effectiveness of a lockdown on air quality. We find that NO_2_ column quantities started following a decreasing trend during the last week of February in Italy and US which indicate a partial unofficial closure of anthropogenic activities, taking into consideration that the official COVID-19 induced lockdown was enforced on 09 March and around 25 March in Italy and the US, respectively. Whereas, in the UK, an increasing trend in NO_2_ column till 10 May indicates no such public awareness to restrict anthropogenic activities (the government declared the lockdown from 23 March). We acknowledge that the reduction in NO_2_ is also associated with the compliance of the population of the individual nation to abide by the lockdown measures.

Furthermore, we suggest that the interventions employed by India may not be at the time when the curve is very far from reaching the transition; however, the smaller proportion of affected cases may be the determining factor in limiting the disease spread in India. Implementation of a nationwide lockdown in India may have better prepared the country for taking measures to control the epidemic spread and bend the curve. However, our analysis also suggests that the period beyond the signals of CSD also needs efficient monitoring. The result of our minimal stochastic model predicts that on 20 May, the number of infected people can go up to approximately 1,09,262. Thus, the extended period of such measures are needed and likely to be effective [6].

We envision that it is fundamental to identify the situation of such a crisis across the world and make use of the lead time. The EWSs can keep track of the changes in the trend statistics in the number of reported cases and warn when a threshold is reached. The statistical tools used can be beneficial to identify whether the features of shift in a system are suppressed by the intervention strategies being adopted. In particular, while different combinations of strategies are adopted to overcome such crisis, the information of an upcoming transition and its threshold is important to formulate the degree of such interventions. However, special care should be taken in the choice of rolling window size and the filtering bandwidth while estimating the signals of slowing down. Inappropriate choices may give weak and/or diminish the signals of an imminent transition, which may deviate from understanding the urgency of the situation. Another aspect to consider is that the varying extent of testing for COVID-19 across the countries may have affected the total number of reported cases; thus, our results here hold specifically for the number of reported cases.

## MATERIALS AND METHODS

### The COVID-19 Data Source

We have used the COVID-19 data set provided by the European Centre for Disease Prevention and Control (ECDC): An agency of the European Union (available from https://www.ecdc.europa.eu/en/publicationsdata). Initially we extract the data of the daily number of reported cases up to 25 March, 2020 and in general mark the first date of the reported cases as the day of the beginning of the epidemic in the respective countries. Regardless of the affected person recovers or dies, the virus contraction occurs once; thus, we consider cumulative data of the daily number of the confirmed cases for nine different countries for our study.

### Data selection

We use the available time-series to test the predictability of an upcoming transition for each country. The generic indicators are examined using the time-series segments before the transition in the number of cases of the epidemic (see *SI Appendix, section 4.: Detection of the Transition Threshold*) in each country (shaded regions in Fig. 1).

### Detrending

Often, non-stationarities in the data lead to false indications of impending transitions. To overcome this, we obtain the residual time-series by subtracting a Gaussian kernel smoothing function from the empirical time-series [23]. Further, we estimate the variance and autocorrelation at first lag for the residual time-series choosing a rolling window size from the sensitivity analyses of the time-series data for each country. We choose the filtering bandwidth avoid any under-fit or over-fit (for details see *SI Appendix, Table S2*).

### Autocorrelation at first lag and variance

The fluctuations in the time-series reveal different novel phenomena such as sudden transition, flickering and stochastic switching, etc. It is established that followed by a perturbation, the rate of return of the system slows down near an impending transition or a tipping point. This phenomenon of slow return rate or recovery from a perturbation in the vicinity of a sudden transition is known as critical slowing down (CSD). We capture the signals of CSD by estimating changes in the short term autocorrelation (at lag-1) and variance of the time-series. CSD increases the short term memory of the time-series, which is observed through the correlation structure of the time-series before a transition. We compute auto-correlation at lag-1 by fitting an autoregressive model of order 1 (of the form *z_t_*_+1_ = *α*_1_*z_t_* + *ϵ_t_*) using an ordinary least-squares fitting method. The time-series analysis has been performed using the “Early Warning Signals Tool-box” (http://www.early-warning-signals.org/).

### Sensitivity analysis

The predictability of each of the indicator depends upon the data sets investigated as well as the choices made for processing the data. Thus, it is essential to check the efficacy of our results to such choices. In particular, we analyse the sensitivity of our observations to the choice of rolling window size and degree of smoothing (filtering bandwidth) used during the calculation of indicators and detrending/filtering the data sets, respectively. We estimate the CSD indicators using window sizes ranging from 40% to 90% of the time-series length in an increment of 1 point and for bandwidths ranging from 5% to 100% with the increment of 1 point. We quantify the robustness of the outcomes towards the range of window sizes and bandwidth using the distribution of the Kendall-*τ* test statistic.

### Surrogates

To test the significance of our statistical analysis, we estimate Kendall rank correlation-*τ* test statistic for both the generic indicators. We generate 1000 surrogate time-series of the same length as the analysed real data sets to test the likelihood of obtaining the computed trends by chance. The surrogate records are obtained on bootstrapping the real data sets by shuffling the original residual time-series and sampling the data with replacement. This method generates the surrogate time-series with a similar distribution of the original time-series [27]. For each surrogate, we consider the Kendall-*τ* estimate as the test statistic to measure the robustness of the outcomes. Further, we calculate the fraction of the surrogates having the same or higher test static value than the original data and measure the probability *P* (*τ*^*^ ≤ *τ*) to calculate that the observed test statistic is by chance. We also generate surrogate time-series using phase randomisation method (for details see *SI Appendix, section 2.: Surrogate analysis*).

### Satellite retrieved total column NO_2_

Worldwide, the lockdown response to the onset and spread of COVID-19 caused a decrease in daily and economic activities, which in turn is expected to cause a reduction in ambient air pollution. This can also be used as an indicator to determine whether government policies of lockdowns/restricted human movements are successful or not. To further examine this, we use the Ozone Monitoring Instrument (OMI) retrieved total column NO_2_ (available from https://aura.gsfc.nasa.gov/omi.html) as a proxy to infer the change in anthropogenic air pollution for the time-period of our study. OMI flies onboard the EOS Aura sun-synchronous polar-orbiting satellite. It has a swath length of 2600km and a level-2, spatial resolution of 13×24km^2^ [48]. The OMI NO_2_ column was satisfactorily validated against surface spectrometer measurements in recent studies [49, 50]. To roughly obtain a global coverage, we consider 3-day time slices (triads) within which the overlapping swath overpasses were averaged. Thereafter, we perform a population-weighted average of the grids that lie within the political boundaries of the countries considered in this study. Gridded population data was obtained for 2015 from SEDAC (https://sedac.ciesin.columbia.edu/data/collection/gpwv4).

## Data Availability

The COVID-19 Data Source. We have used the COVID-19 data set provided by the European Centre for Disease Prevention and Control (ECDC): An agency of the European Union (https://www.ecdc.europa.eu/en/publications-data/download-todays-data-geographic-distribution-covid-19-cases-worldwide). (The data is freely available and we have cited it properly in the paper).
To further examine this, we use the Ozone Monitoring Instrument (OMI) retrieved total column NO2 (available from https://aura.gsfc.nasa.gov/omi.html) as a proxy to infer the change in anthropogenic air pollution for the time period of our study.

https://www.ecdc.europa.eu/en/publications-data/download-todays-data-geographic-distribution-covid-19-cases-worldwide

https://aura.gsfc.nasa.gov/omi.html

## ACKNOWLEDGMENTS

S.S. acknowledges the financial support from the Department of Science & Technology (DST), Govt. of India under the scheme DST-Inspire [Grant No.: IF160459]. P.S.D. acknowledges financial support from the Science & Engineering Research Board (SERB), Govt. of India [Grant No.: CRG/2019/002402]. This manuscript has been released as a pre-print at medRxiv, (Kaur et al. [51]).

## References

[1] World Health Organization. Coronavirus disease 2019 (covid-19): situation report, 59. Technical report, World Health Organization, 2020.

[2] Eurosurveillance editorial team. Note from the editors: World health organization declares novel coronavirus (2019-ncov) sixth public health emergency of international concern. Eurosurveillance, 25(5), 2020.

[3] Covid-19 coronavirus pandemic. https://www.worldometers.info/coronavirus/, 2020.

[4] Joseph T Wu, Kathy Leung, Mary Bushman, Nishant Kishore, Rene Niehus, Pablo M de Salazar, Benjamin J Cowling, Marc Lipsitch, and Gabriel M Leung. Estimating clinical severity of covid-19 from the transmission dynamics in wuhan, china. Nature Medicine, pages 1–5, 2020.

[5] Moritz U. G. Kraemer, Chia-Hung Yang, Bernardo Gutierrez, Chieh-Hsi Wu, Brennan Klein, David M. Pigott, Louis du Plessis, Nuno R. Faria, Ruoran Li, William P. Hanage, John S. Brownstein, Maylis Layan, Alessandro Vespignani, Huaiyu Tian, Christopher Dye, Oliver G. Pybus, and Samuel V. Scarpino. The effect of human mobility and control measures on the covid-19 epidemic in china. Science, 2020.

[6] Rajesh Singh and R Adhikari. Age-structured impact of social distancing on the covid-19 epidemic in india. *arXiv preprint arXiv:2003.12055*, 2020.

[7] Elisabeth Mahase. Covid-19: UK starts social distancing after new model points to 260 000 potential deaths, 2020.

[8] Coronavirus in new york: Lunar new year events canceled over fears. https://www.nytimes.com/2020/01/29/nyregion/coronavirus-nyc.html, 2020.

[9] Richard J Hatchett, Carter E Mecher, and Marc Lipsitch. Public health interventions and epidemic intensity during the 1918 influenza pandemic. Proceedings of the National Academy of Sciences U.S.A., 104(18):7582–7587, 2007.

[10] Martin CJ Bootsma and Neil M Ferguson. The effect of public health measures on the 1918 influenza pandemic in us cities. Proceedings of the National Academy of Sciences U.S.A., 104(18):7588–7593, 2007.

[11] Neil Ferguson, Daniel Laydon, Gemma Nedjati Gilani, Natsuko Imai, Kylie Ainslie, Marc Baguelin, Sangeeta Bhatia, Adhiratha Boonyasiri, ZULMA Cucunuba Perez, Gina Cuomo-Dannenburg, et al. Report 9: Impact of non-pharmaceutical interventions (npis) to reduce covid19 mortality and healthcare demand. Technical report, Imperial College London, 2020.

[12] Roy M Anderson, Hans Heesterbeek, Don Klinkenberg, and T Déirdre Hollingsworth. How will country-based mitigation measures influence the course of the covid-19 epidemic? The Lancet, 395(10228):931–934, 2020.

[13] Kiesha Prem, Yang Liu, Timothy W Russell, Adam J Kucharski, Rosalind M Eggo, Nicholas Davies, Stefan Flasche, Samuel Clifford, Carl A B Pearson, James D Munday, Sam Abbott, Hamish Gibbs, Alicia Rosello, Billy J Quilty, Thibaut Jombart, Fiona Sun, Charlie Diamond, Amy Gimma, Kevin [van Zandvoort], Sebastian Funk, Christopher I Jarvis, W John Edmunds, Nikos I Bosse, Joel Hellewell, Mark Jit, and Petra Klepac. The effect of control strategies to reduce social mixing on outcomes of the covid-19 epidemic in wuhan, china: a modelling study. The Lancet Public Health, 2020.

[14] Suzanne M O’Regan and John M Drake. Theory of early warning signals of disease emergence and leading indicators of elimination. Theoretical Ecology, 6(3):333–357, 2013.

[15] Tobias S Brett, John M Drake, and Pejman Rohani. Anticipating the emergence of infectious diseases. Journal of The Royal Society Interface, 14(132):20170115, 2017.

[16] Eamon B O’Dea and John M Drake. Disentangling reporting and disease transmission. Theoretical Ecology, 12(1):89–98, 2019.

[17] John M Drake, Tobias S Brett, Shiyang Chen, Bogdan I Epureanu, Matthew J Ferrari, Éric Marty, Paige B Miller, Eamon B O’Dea, Suzanne M O’Regan, Andrew W Park, et al. The statistics of epidemic transitions. PLoS Computational Biology, 15(5), 2019.

[18] Mallory J Harris, Simon I Hay, and John M Drake. Early warning signals of malaria resurgence in kericho, kenya. Biology Letters, 16(3):20190713, 2020.

[19] Sonia Kefi, Vasilis Dakos, Marten Scheffer, Egbert H. Van Nes, and Max Rietkerk. Early warning signals also precede non-catastrophic transitions. Oikos, 122(5):641–648, 2013.

[20] Ying Liu, Albert A Gayle, Annelies Wilder-Smith, and Joacim Rocklv. The reproductive number of COVID19 is higher compared to SARS coronavirus. Journal of Travel Medicine, 27(2), 02 2020. taaa021.

[21] Christopher J Dibble, Eamon B O’Dea, Andrew W Park, and John M Drake. Waiting time to infectious disease emergence. Journal of The Royal Society Interface, 13(123):20160540, 2016.

[22] M. Scheffer, S. R. Carpenter, T. M. Lenton, J. Bascompte, W. A. Brock, V. Dakos, J. van de Koppel, I. A. van de Leemput, S. A. Levin, E. H. van Nes, M. Pascual, and J. Vandermeer. Anticipating critical transitions. Science, 338:344–348, 2012.

[23] V. Dakos, S. R. Carpenter, W. A. Brock, A. M. Ellison, V. Guttal, A. R. Ives, S. Kéfi, V. Livina, D. A. Seekell, E. H. van Nes, and M. Scheffer. Methods for Detecting Early Warnings of Critical Transitions in Time Series Illustrated Using Simulated Ecological Data. PLoS One, 7:e41010, 2012.

[24] Rong Wang, John A Dearing, Peter G Langdon, Enlou Zhang, Xiangdong Yang, Vasilis Dakos, and Marten Scheffer. Flickering gives early warning signals of a critical transition to a eutrophic lake state. Nature, 492(7429):419, 2012.

[25] Marten Scheffer, Jordi Bascompte, William A Brock, Victor Brovkin, Stephen R Carpenter, Vasilis Dakos, Hermann Held, Egbert H Van Nes, Max Rietkerk, and George Sugihara. Early-warning signals for critical transitions. Nature, 461(7260):53–59, 2009.

[26] Sukanta Sarkar, Sudipta Kumar Sinha, Herbert Levine, Mohit Kumar Jolly, and Partha Sharathi Dutta. Anticipating critical transitions in epithelial–hybrid-mesenchymal cell-fate determination. Proceedings of the National Academy of Sciences U.S.A., 116(52):26343–26352, 2019.

[27] Vasilis Dakos, Marten Scheffer, Egbert H van Nes, Victor Brovkin, Vladimir Petoukhov, and Hermann Held. Slowing down as an early warning signal for abrupt climate change. Proceedings of the National Academy of Sciences U.S.A., 105(38):14308–14312, 2008.

[28] M. Scheffer. Critical Transitions in Nature and Society. Princeton University Press, 2009.

[29] R. M. May, S. A. Levin, and G. Sugihara. Complex systems: Ecology for bankers. Nature, 451:893–895, 2008.

[30] Marten Scheffer, Steve Carpenter, Jonathan A Foley, Carl Folke, and Brian Walker. Catastrophic shifts in ecosystems. Nature, 413(6856):591, 2001.

[31] Marten Scheffer, J Elizabeth Bolhuis, Denny Borsboom, Timothy G Buchman, Sanne MW Gijzel, Dave Goulson, Jan E Kammenga, Bas Kemp, Ingrid A van de Leemput, Simon Levin, et al. Quantifying resilience of humans and other animals. Proceedings of the National Academy of Sciences U.S.A., 115(47):11883–11890, 2018.

[32] Vasilis Dakos, Egbert H Van Nes, Paolo d’Odorico, and Marten Scheffer. Robustness of variance and autocorrelation as indicators of critical slowing down. Ecology, 93(2):264–271, 2012.

[33] Partha Sharathi Dutta, Yogita Sharma, and Karen C Abbott. Robustness of early warning signals for catastrophic and non-catastrophic transitions. Oikos, 127(9):1251–1263, 2018.

[34] Suzanne M O?Regan and John M Drake. Theory of early warning signals of disease emergenceand leading indicators of elimination. Theoretical Ecology, 6(3):333–357, 2013.

[35] Tobias Brett, Marco Ajelli, Quan-Hui Liu, Mary G Krauland, John J Grefenstette, Willem G van Panhuis, Alessandro Vespignani, John M Drake, and Pejman Rohani. Detecting critical slowing down in high-dimensional epidemiological systems. PLoS computational biology, 16(3):e1007679, 2020.

[36] Wei-jie Guan, Wen-hua Liang, Yi Zhao, Heng-rui Liang, Zi-sheng Chen, Yi-min Li, Xiao-qing Liu, Ru-chong Chen, Chun-li Tang, Tao Wang, Chun-quan Ou, Li Li, Ping-yan Chen, Ling Sang, Wei Wang, Jian-fu Li, Caichen Li, Li-min Ou, Bo Cheng, Shan Xiong, Zheng-yi Ni, Jie Xiang, Yu Hu, Lei Liu, Hong Shan, Chun-liang Lei, Yi-xiang Peng, Li Wei, Yong Liu, Ya-hua Hu, Peng Peng, Jian-ming Wang, Ji-yang Liu, Zhong Chen, Gang Li, Zhi-jian Zheng, Shao-qin Qiu, Jie Luo, Chang-jiang Ye, Shao-yong Zhu, Lin-ling Cheng, Feng Ye, Shi-yue Li, Jin-ping Zheng, Nuo-fu Zhang, Nan-shan Zhong, and Jian-xing He. Comorbidity and its impact on 1590 patients with covid-19 in china: A nationwide analysis. European Respiratory Journal, 2020.

[37] Servio Pontes Ribeiro, Wesley Dattilo, Alcides Castro e Silva, Alexandre Barbosa Reis, Aristoteles Goes-Neto, Luiz Alcantara, Marta Giovanetti, Geraldo Wilson Fernandes, Vasco Ariston Azevedo, and Wendel Coura-vital. Severe airport sanitarian control could slow down the spreading of covid-19 pandemics in brazil. medRxiv, 2020.

[38] Joan T Matamalas, Sergio Gómez, and Alex Arenas. Abrupt phase transition of epidemic spreading in simplicial complexes. Physical Review Research, 2(1):012049, 2020.

[39] Natalie M Linton, Tetsuro Kobayashi, Yichi Yang, Katsuma Hayashi, Andrei R Akhmetzhanov, Sung-mok Jung, Baoyin Yuan, Ryo Kinoshita, and Hiroshi Nishiura. Incubation period and other epidemiological characteristics of 2019 novel coronavirus infections with right truncation: a statistical analysis of publicly available case data. Journal of Clinical Medicine, 9(2):538, 2020.

[40] Ruiyun Li, Sen Pei, Bin Chen, Yimeng Song, Tao Zhang, Wan Yang, and Jeffrey Shaman. Substantial undocumented infection facilitates the rapid dissemination of novel coronavirus (SARS-CoV2). Science, 2020.

[41] Zander S. Venter, Kristin Aunan, Sourangsu Chowdhury, and Jos Lelieveld. Covid-19 lockdowns cause global air pollution declines. Proceedings of the National Academy of Sciences U.S.A., 2020.

[42] Daniel J Jacob and Darrell A Winner. Effect of climate change on air quality. Atmospheric environment, 43(1):51–63, 2009.

[43] Daniel T Gillespie. Stochastic simulation of chemical kinetics. Annu. Rev. Phys. Chem., 58:35–55, 2007.

[44] Jodie Lynn Guest, Carlos del Rio., and Travis Sanchez. The 3 steps needed to end the covid-19 pandemic: Bold public health leadership, rapid innovations, and courageous political will. JMIR Public Health and Surveillance, 2020.

[45] Coronavirus disease 2019 (covid-19) situation report 72. https://www.who.int/docs/default-source/coronaviruse/situation-reports/20200401-sitrep-72-covid-19.pdf?sfvrsn=3dd8971b_2, 2020.

[46] Coronavirus disease 2019 (covid-19) situation report 74. https://www.who.int/docs/default-source/coronaviruse/situation-reports/20200403-sitrep-74-covid-19-mp.pdf?sfvrsn=4e043d03_14, 2020.

[47] Coronavirus disease 2019 (covid-19) situation report 76. https://www.who.int/docs/default-source/coronaviruse/situation-reports/20200405-sitrep-76-covid-19.pdf?sfvrsn=6ecf0977_2, 2020.

[48] Nickolay A. Krotkov, Lok N. Lamsal, Edward A. Celarier, William H. Swartz, Sergey V. Marchenko, Eric J. Bucsela, Ka Lok Chan, Mark Wenig, and Marina Zara. The version 3 OMI NO_2_ standard product. Atmospheric Measurement Techniques, 10(9):3133–3149, September 2017.

[49] Yang Wang, Steffen Beirle, Johannes Lampel, Mariliza Koukouli, Isabelle De Smedt, Nicolas Theys, Ang Li, Dexia Wu, Pinhua Xie, Cheng Liu, Michel Van Roozendael, Trissevgeni Stavrakou, Jean-François Müller, and Thomas Wagner. Validation of OMI, GOME-2A and GOME-2B tropospheric NO_2_, SO_2_ and HCHO products using MAX-DOAS observations from 2011 to 2014 in Wuxi, China: investigation of the effects of priori profiles and aerosols on the satellite products. Atmospheric Chemistry & Physics, 17(8):5007–5033, 2017.

[50] LN Lamsal, NA Krotkov, EA Celarier, WH Swartz, KE Pickering, EJ Bucsela, JF Gleason, RV Martin, S Philip, H Irie, et al. Evaluation of omi operational standard no2 column retrievals using in situ and surface-based no2 observations. Atmospheric Chemistry and Physics, 14(21):11587, 2014.

[51] Taranjot Kaur, Sukanta Sarkar, Sourangsu Chowdhury, Sudipta Kumar Sinha, Mohit Kumar Jolly, and Partha Sharathi Dutta. Anticipating the novel coronavirus disease (covid-19) pandemic. *medRxiv*, 2020.

